# Virus shedding patterns in nasopharyngeal and fecal specimens of COVID-19 patients

**DOI:** 10.1101/2020.03.28.20043059

**Authors:** Ning Zhang, Yuhuan Gong, Fanping Meng, Yuhai Bi, Penghui Yang, Fusheng Wang

## Abstract

Diagnosis is the key point for confirmation and treatment of COVID-19. we focused on comparative analysis of virus dynamics between the upper respiratory and feces specimens in the COVID-19 patients. A total of 66 upper respiratory swabs, 51 feces, 56 urine and 56 plasma samples were sequentially collected from 23 patients in a designated hospital. The plasma and urine samples were all negative, except for urine samples from two severe cases at the latest available detection point. Conversely, virus was shed in respiratory swabs and feces samples during the diseased period. Ten of 12 (83.3%) cases were positive for feces samples, while 14 of 21 (66.7%) were positive for respiratory samples. In addition, the median duration of virus shedding was 10.0 days (IQR 8.0 to 17.0) in the upper respiratory swabs, but was 22.0 days (IQR 15.5 to 23.5) for the feces. Notably, at 26 days after discharge, case 3 (a 45-year-old) was detected positive again in the feces samples, but appears to be healthy and negative for respiratory swabs. These results indicated that beside respiratory samples, intestinal samples (e.g. feces) should be recommended for diagnosis of COVID-19, especially before a patient discharge and for monitoring the relapse of discharged patients.

---

An acute viral pneumonia (COVID-19), caused by the novel coronavirus HCoV-19, was first identified during December 2019 in China(1). HCoV-19 was found to be highly transmissible in humans(2) and is now a pandemic, with transmission into over 140 countries, causing over 150,000 infections and 6,000 deaths as of March 15, 2020 (3).

Diagnosis is critical for confirmation and treatment of COVID-19. Viral RNA detection for the respiratory samples is currently the primary criteria for diagnosis of COVID-19. Two studies on virus loads in clinical samples have been recently reported, in which viral loads in nasal and throat swabs and sputum specimens peaked at three to seven days after illness onset (d.a.o.) and virtually disappeared before 15 d.a.o.(4, 5). Another study showed that the median duration of virus shedding in throat swabs was 20 d.a.o. in survivors and was detectable until death in non-survivors(6). Additionally, live viruses have been isolated in the feces and urine samples of COVID-19 patients. However, the viral dynamics in these two types of specimens have not yet been elucidated, as well as comparative studies on virus shedding in the upper respiratory, intestinal and urinary tracts.

From January 20 to February 23, 2020, a total of 23 patients were treated in a designated hospital in Beijing (11 were imported, 12 were secondary cases; two family clusters; 12 men, 11 women; median age was 48.0 years (IQR 40.0 to 62.0); two with severe disease, the others were mild-to-moderate, all patients recovered except for one due to an unrelated bacterial infection) (Table S1 in the Supplementary appendix). Upper respiratory (nasal-throat mixed) swabs (n= 66), feces (n= 51), urine (n= 56), and plasma (n= 56) samples were collected for viral RNA detection by real-time RT-PCR (rRT-PCR). The study was approved by the Ethics Committees of Chinese Academy of Sciences (SQIMCAS20). Informed consent was obtained from all subjects for being included in the study, and all patient data were anonymized before study inclusion.

The plasma and urine samples were all negative, except for urine samples from two severe cases at the latest available detection point (16 or 21 d.a.o). Conversely, virus was shed in respiratory swabs and feces samples during the diseased period (Figure 1A). Ten of 12 cases (83.3%) were positive for feces samples, while 14 of 21 cases (66.7%) were positive for respiratory samples. In addition, all samples from one severe patient were negative until 21 d.a.o., when feces samples were positive. The median duration of virus shedding was 10.0 days (IQR 8.0 to 17.0) in nasal-throat mixed swabs, but was 22.0 days (IQR 15.5 to 23.5) for the feces (Figure 1 B). The viral titers of nasal-throat swabs peaked at six to nine d.a.o. and at 14-18 d.a.o for fecal samples, and the highest virus titers at the peak was significantly higher for feces (10^5.8^ copies/ml, mean 5623 copies/ml) than of respiratory samples (10^6.3^ copies/ml, mean 2535 copies/ml) (Figure 1A). Notably, at 26 days after discharge, case 3 was detected positive again in feces samples, but appears to be healthy and negative for respiratory swabs. These results indicated that beside respiratory samples, intestinal samples (e.g. feces) should be recommended for diagnosis of COVID-19, especially for monitoring the relapse of discharged patients.

**Figure 1.**
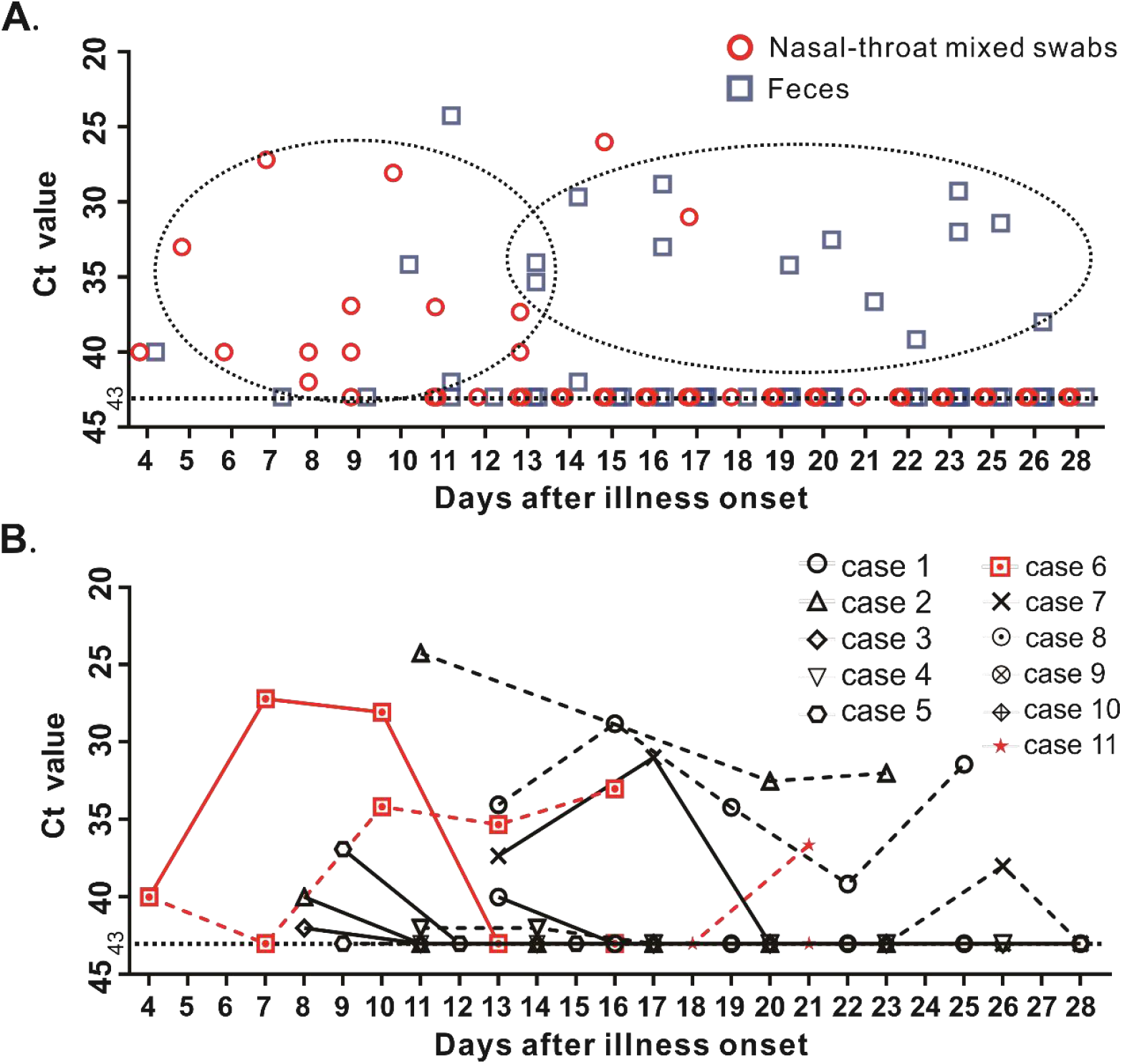
Virus dynamics in nasopharyngeal and fecal specimens of COVID-19 cases. The nasal-throat mixed swabs and fecal samples of all 23 cases were detected by rRT-PCR targeting ORF1ab, N and S genes (Mabsky Biotech Co., Ltd., CONFORMITE EUROPEENNE NO.), the Ct values of ORF1ab gene were shown in (A). Sequential nasal-throat mixed swabs, feces, urine, and plasma samples were collected in 11 cases and used for virus detection. The Ct values of nasal-throat mixed swabs (solid line) and feces (dotted line) specimens were shown in (B). Two cases with severe diseases, identified according to the guideline of HCoV-19 infection from the National Health Commission of the People’s Republic of China, were colored in red. The other eight cases with mild-to-moderate symptoms were in black. Ct values were inversely related to viral RNA copy number, with Ct values of 37.6, 32.64, 29.22, and 25.77 corresponding to 1×10^3^, 1×10^4^, 1×10^5^, and 1×10^6^ copies per mL. Negative samples were denoted with a Ct value of 43, which was the limit of detection.

We declare no competing interests. This work was supported by the Strategic Priority Research Program of the Chinese Academy of Sciences (CAS) (Grant No. XDB29010102). Y.B. is supported by the NSFC Outstanding Young Scholars (Grant No. 31822055), and Youth Innovation Promotion Association of CAS (Grant No. 2017122). We declare no competing interests.

## Data Availability

We promise all data referred to in the manuscript is availability.

